# Increasing Virus Test Capacity via Recursive Pool Testing with an Application to SARS-CoV-2 Testing

**DOI:** 10.1101/2020.07.02.20144956

**Authors:** Petra Mutzel, Alexander Bertram, Paul Jünger, Pauline Jünger, Horst Krieger, Stephan Schmitz, Michael Jünger

## Abstract

In the context of adequate reactions to the current Covid-19 pandemic, Seifried, Ciesek et al. [6, 5] have proposed the application of SARS-CoV-2 pool testing in the pursuit of increasing testing capacity. We show how this method can be substantially improved in realistic scenarios, and we point out a possible impact on the ongoing discussion concerning the need of increased testing as a complementary measure to relaxed restrictions.

## 1 Introduction

Seifried, Ciesek, et al. [6, 5] have proposed the application of SARS-CoV-2 pool testing in April 2020, see also [2, 1]. The described procedure uses two aliquots of each sample. The first set of aliquots is partitioned into pools of a given size and these pools are tested. A negative test result means that all corresponding samples in the pool are negative, a positive test result requires testing the corresponding samples in the pool individually using the corresponding second set of aliquots.

The authors state: “Dabei wird der Abstrichtupfer zunächst in ein Archivröhrchen gegeben und anschließend in ein Poolgefäß. Da sich bei dieser Poolmethode das Volumen im Poolgefäß nicht vermehrt, wird auch keine Verdünnung und damit keine Abnahme der Empfindlichkeit (Sensitivität) beobachtet.”^1^

According to the article, this pool test procedure was developed and patented by the Goethe University and the DRK blood donation service and leads to the fact that a constantly demanded extension of testing is made possible. The procedure was tested on 50 samples, of which 5 were SARS-CoV-2 positive. These 50 samples were divided into 10 pools of 5 samples each.

In a newer article, Lohse et al. [4] report on testing 1,191 samples, 23 of which are positive, they used pools of size 30 that were again subdivided into subpools of size 10, and they needed only 267 tests. This strategy requires 3 (rather than 2) aliquots of each sample. The authors state that borderline samples might escape detection in pools of size 30. However, they argue that those stem from almost recovered persons.

Both research groups emphasize a tremendously increased testing capacity when the infection rate is low and consequently, many pool tests will have a negative result.

We review the method in [6, 5], henceforth called “the Frankfurt method”, also in view of the variant in [4], henceforth called “the Saarbrücken variant”, propose a new variant based on recursive pool testing, demonstrate its potential in comparison to the Frankfurt method as well as the Saarbruücken variant, and discuss the possible impact on strategies that accompany the current relaxation of restrictions due to the Covid-19 pandemic.

## 2 Pool testing methods

The idea is to combine many samples into one combined sample (“pool”) and apply the test to the combined sample. If the result is negative, all samples in the combined sample are negative. If the result is positive, further testing is necessary in order to determine negativity/positivity for all samples in the combined sample. This idea has already been applied for syphilis testing of soldiers in the 1940s, and is popular for HIV testing since 2009.

### 2.1 The Frankfurt pool testing method

We are given n samples {*s*_1_, *s*_2_, …, *s_n_*} for testing. The Frankfurt pool testing method proceeds as follows:

1. Prepare to divide each sample *s_i_* into up to 2 aliquots *s_i_*,_1_ and *s_i_*,_2_.
2. Partition the set of all *n* samples into subsets of a given size *p* < *n*. (In [6] we have *n* = 50 and *p* = 5, in [4] we have *n* = 1,191 and *p* = 30.)
3. Make a pool test for each subset using the aliquots *s_i_*,_1_. (These tests can be performed simultaneously.)
4. Declare all samples within each negatively tested subset negative.
5. Test all samples within each positively tested subset individually using the aliquots *s_i_*_,2_.

### 2.2 Our recursive pool testing method

Rather than testing all samples in a positive pool, the idea is to again split this pool subset into 2 (or more) smaller subsets for pool testing, and keep splitting as long as possible.

In detail, our enhancement concerns steps 1 and 5 of the Frankfurt pool testing method. For ease of exposition, we assume *p* to be a power of 2, i.e., *p* = 2*^k^* for some *k*. (The method can be easily adapted to arbitrary values of *p*.) In step 1, we moderately increase the number of aliquots:

1’. Prepare to divide each sample *s_i_* into up to *a* = 1 + log_2_ *p* aliquots {*s_i,_*_1_*,s_i,_*_2_, …, *s_i,a_*}.

Step 5 is replaced by an application of the *Divide&Conquer* principle that is well-known in Computer Science:

5’. For each positively tested subset apply the divide&conquer method decribed next.

### Divide&conquer method

For ease of exposition, we assume the pool subset to consist of the sample aliquots

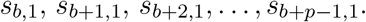

These are the *p* consecutive aliquots starting at index *b*. E.g., for *n* = 16 and *p* = 4, the pool subsets are

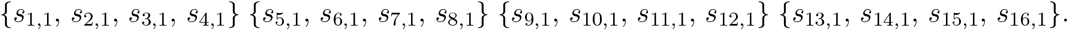

Then, with *e* = *b* + *p* − 1, *dc_test*(*b*, *e*, *d*) stands for “Test the pool of the samples *s_b_*, *s_b_*_+1_, *s_b_*_+2_, …, *s_b_*_+_*_p_*_−1_ using aliquots *d*”.

So our task is *dc_test*(*b*, *e*, 1). Here is *dc_test*(*b*, *e*, *d*) in pseudocode:

*de_test*(*b*, *e*, *d*)

**if** *b* = *e* **then**

Perform an individual test with sample aliquot *s_b,d_*.

**if** the test is negative **then**

Declare sample *s_b_* negative.

**else**

Declare sample *s_b_* positive.

**end if**

**else**

Perform a pool test with the sample aliquots *s_b,d_*, *s_b_*_+1_*_,d_*, … *s_e,d_*.

**if** the test is negative **then**

Declare all samples *s_b_*, *s_b_*_+1_, …, *s_e_* negative.

**else**

Perform the pool test 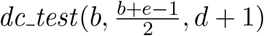.

Perform the pool test 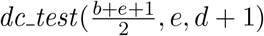.

// The two test above can be performed simultaneously.

**end if**

**end if**

### 2.3 A small example for illustration

We have *n* = 16 samples *s*_1_, *s*_2_, …, *s*_16_ all of which are negative but two, namely *s*_5_ and *s*_15_, which are positive. The pool size is *p* = 4.

Both methods apply pool tests to the subsets

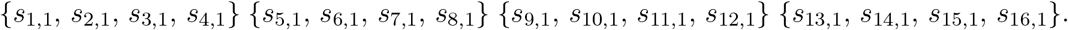

The first and the third will have a negative result, the second and the fourth a positive result. Up to this point, both methods will have applied 4 tests.

The Frankfurt pool testing method will now apply individual tests to the sample aliquots *s*_5,2_, *s*_6,2_, *s*_7,2_, *s*_8,2_, *s*_13,2_, *s*_14,2_, *s*_15,2_, and *s*_16,2_ (8 individual tests) so that a total of 12 tests is performed.

The recursive pool testing method will replace {*s*_5,1_, *s*_6,1_, *s*_7,1_, *s*_8,1_} by {*s*_5,2_, *s*_6,2_} and {*s*_7,2_, *s*_8,2_} and make pool tests for both. It will get a positive result for the first and therefore test *s*_5,3_ and *s*_6,3_ individually. Then it will replace {*s*_13,1_, *s*_14,1_, *s*_15,1_, *s*_16,1_} by {*s*_13,2_, *s*_14,2_} and {*s*_15,2_, *s*_16,2_}, make pool tests for both and get a negative result for the first and a positive result for the second. Therefore it will test *s*_15,3_ and *s*_16,3_ individually. This involves another 4 pool tests plus 4 individual tests, so, again, a total of 12 tests, no improvement in terms of the number of tests.

Here is an illustration of the actions of both methods (boxes correspond to tests, blue means “negative”, red means “positive”. Both methods initially divide the entire set of 16 samples

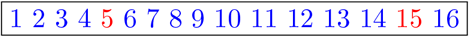

into intervals of 4 sample aliquots

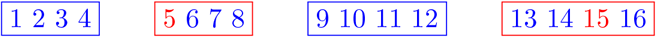

of 4 samples each that are pool tested: The frame colors indicate negative and positive test results.

Afterwards the Frankfurt pool testing method performs the individual tests

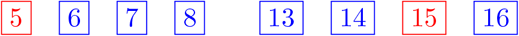

while the recursive method proceeds by performing the pool tests

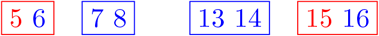

and finally the individual tests

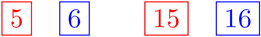

We see that the recursive method requires up to 3 aliquots, namely 1 aliquot for the samples in {*s*_1_, *s*_2_, *s*_3_, *s*_4_, *s*_9_, *s*_10_, *s*_11_, *s*_12_}, 2 aliquots for the samples in {*s*_7_, *s*_8_, *s*_13_, *s*_14_}, and 3 aliquots for the samples in {*s*_5_, *s*_6_, *s*_15_, *s*_16_}.

### 2.4 Generalized recursive method

So far we have split each pool into 2 subpools. Of course, we can just as well split into *B* > 2 subpools. Again, for ease of exposition, we assume that the pool size *p* is a power of *B*, i.e., *p* = *B^k^* for some *k*. In practice, we expect pools in which there is no or only one positive sample. In the former case, no further test is necessary. In the latter case, the positive sample is found after 1 + *B* log*_B_ p* tests. E.g., let *p* = 4 and *B* = 2. Then the pool of size 4 is positive (first test). One of the two subpools of size 2 (second and third test) is positive and requires another two individual tests (fourth and fifth test), i.e., a total of 1 + 2log_2_ 4 = 5 tests. The situation is illustrated in Figure 1. The case *p* = 9 and B = 3 with a total of 1 + 3 log_3_ 9 = 7 tests is illustrated in Figure 2.

**Figure 1:**
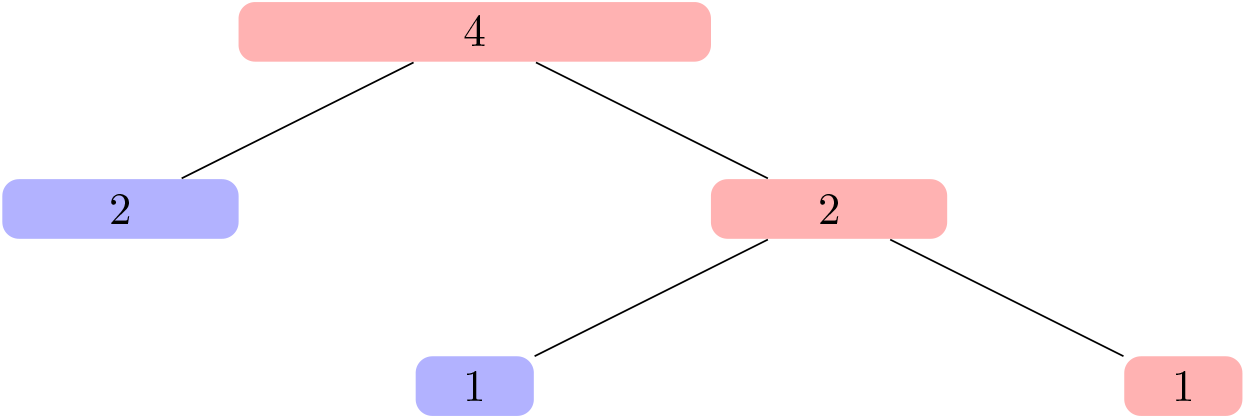
5 tests when *p* = 4 and *B* = 2

**Figure 2:**
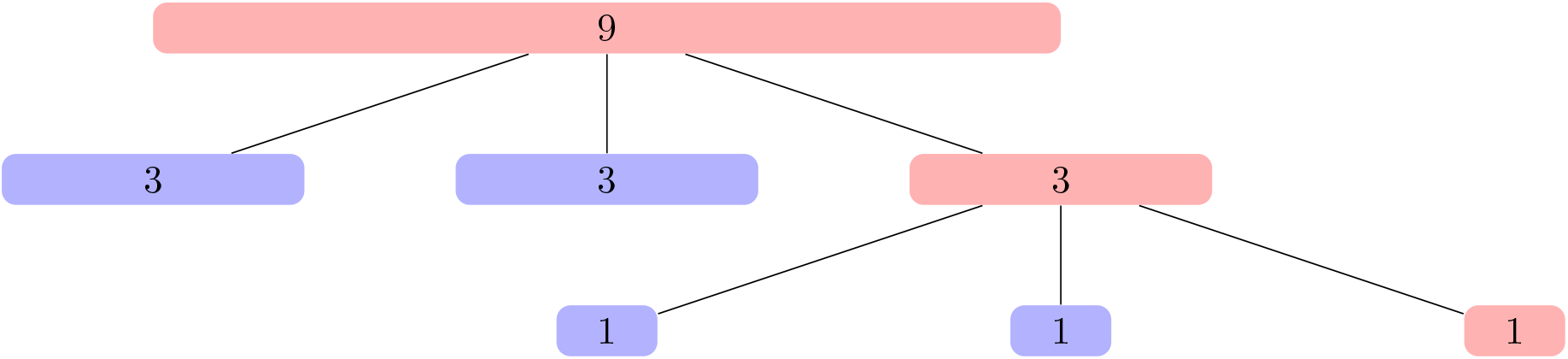
7 tests when *p* = 9 and *B* = 3

What would be the ideal *B* if the pool contains one positive sample? Mathematical calculations show that the unique minimum of *f*_1_(*B,p*) = 1 + *B* log_B_ *p* for *B* ≥ 2 is *B* = *e* = 2.71828… (Euler’s number) for any given pool size *p* ≥ 2. The left picture in Figure 3 shows a plot of the function *f*_1_(*B,p*). But what is the best integer? Since *f*_1_ (2,*p*) > *f*_1_(3,*p*) and *f*_1_ is monotonically increasing with increasing *B* > *e*, we obtain *B* = 3 as the best choice.

**Figure 3:**
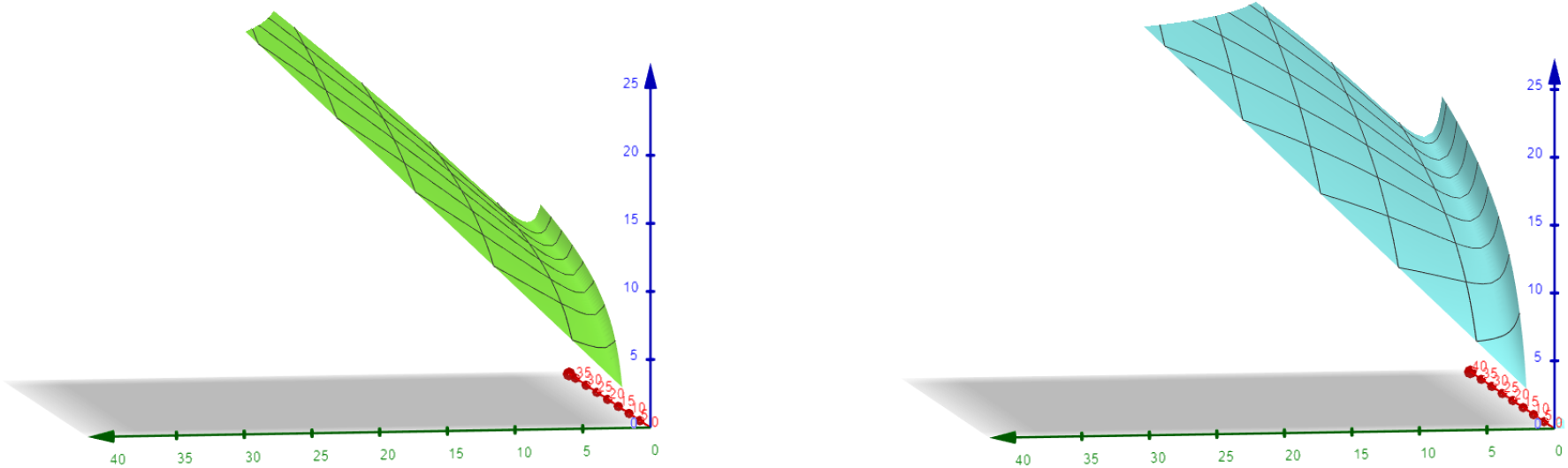
The functions *f*_1_(*B,p*) (left) and *f*_2_(*B,p*) (right). The green axis is associated with *B* and the red axis is associated with *p*. The blue axis shows the expected number of tests for the case of exactly one positive (left) and two positive (right) samples, respectively.

The analysis for two or more positive samples in a pool is more involved. If the pool contains two positive samples, then we can show that the function

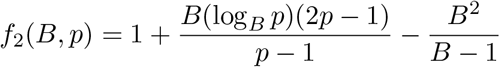

gives the expected number of tests as a function of *B* and *p*. The picture on the right of Figure 3 shows a plot of the function *f*_2_ (*B*,*p*). Again, it turns out that *B* = 3 is the best choice as long as *p* ≥ 13. Indeed, for pool sizes from 7 to 12, it would be optimum to split the pools into 4 pieces, for pool size 6, the value *B* = 5 would be ideal, and smaller pool sizes should be tested individually. Three or more positive samples in a pool of size up to 30 are quite unrealistic. The Appendix contains a detailed mathematical analysis of the above. Our computational tests confirm that *B* = 3 is the best choice in realistic scenarios with pool sizes between 25 and 30. So we restrict our attention in the following to the 3-split version of recursive pool testing.

### 2.5 Statistics for some test simulations

According to the Robert Koch Institute (RKI), the number of tests per week from calendar weeks 2 to 21 varied between 124,000 and 408,000 for about 170 laboratories, which is a total of about 2, 000 tests per laboratory week on average, i.e., 400 per day. The proportion of positive tests in Germany has been between 1.5% and 9%. In the 21st calendar week, 344,782 laboratory tests for the coronavirus (SARS-CoV-2) were carried out in Germany, 5,116 (1.5%) have been positive.

Currently, a typical laboratory seems to test about 400 samples per day. Accordingly, we have simulated tests for *n* = 400 samples with pool size *p* = 27 for varying infection rates. The minima/maxima/averages/medians have been taken over 1,000,000 samples, in which the infected samples are uniformly randomly distributed. The results for the Frankfurt method in comparison to the 3-split version of the recursive method are presented in Table 1 and, for the medians only, as a bar chart in Figure 4. The results show that, with small infection rates up to about 5%, the new approach outperforms the Frankfurt approach by more than 50%. For example, for an infection rate of 1.5%, our method needs about 66 tests compared to the Frankfurt method with 150 tests (which is a reduction of 56%). For larger rates, the advantage is less significant, and for rates from about 25 %, the Frankfurt approach is better.

**Figure 4:**
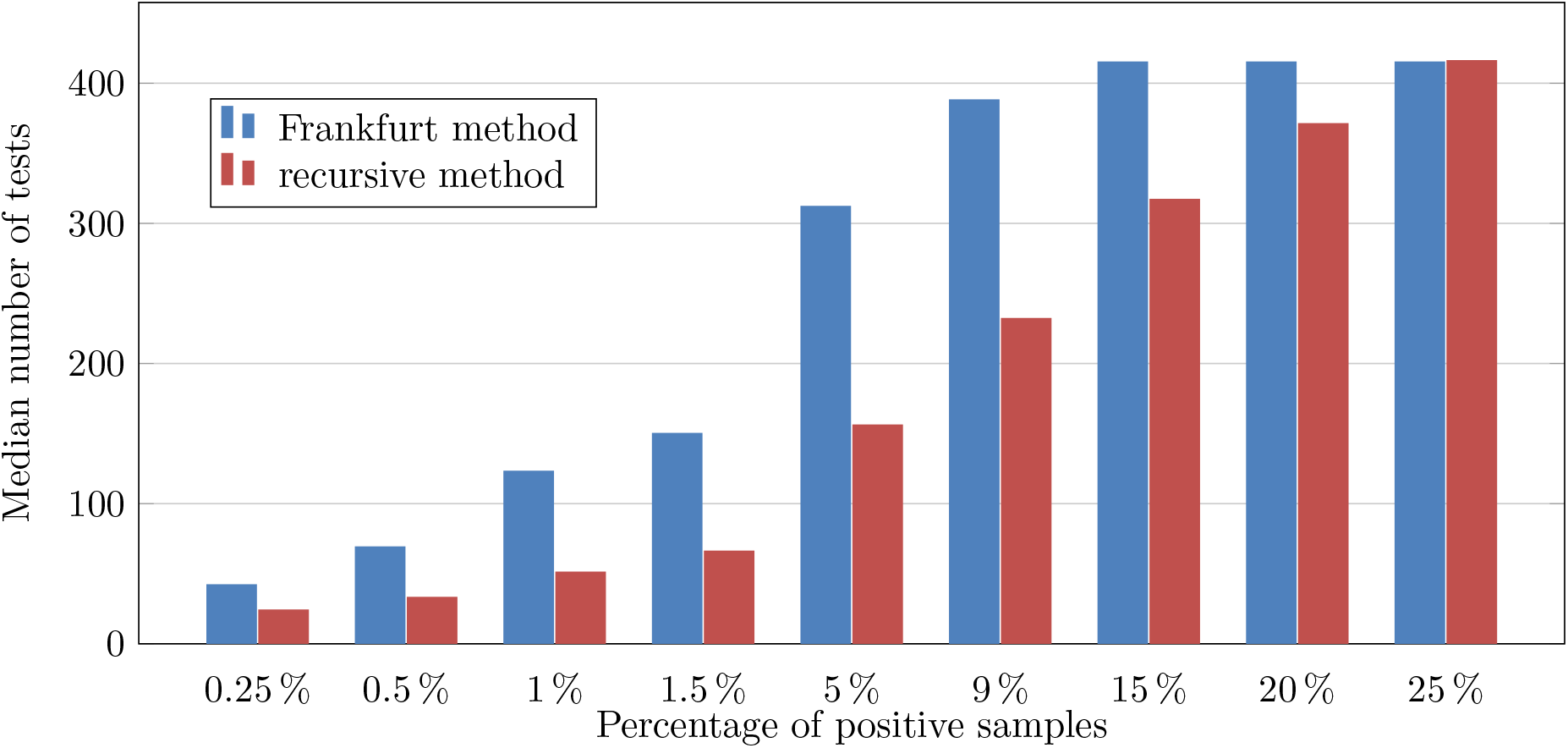
Median number of tests for 400 samples with infection rates from 0.25 % to 25 %

**Figure 5:**
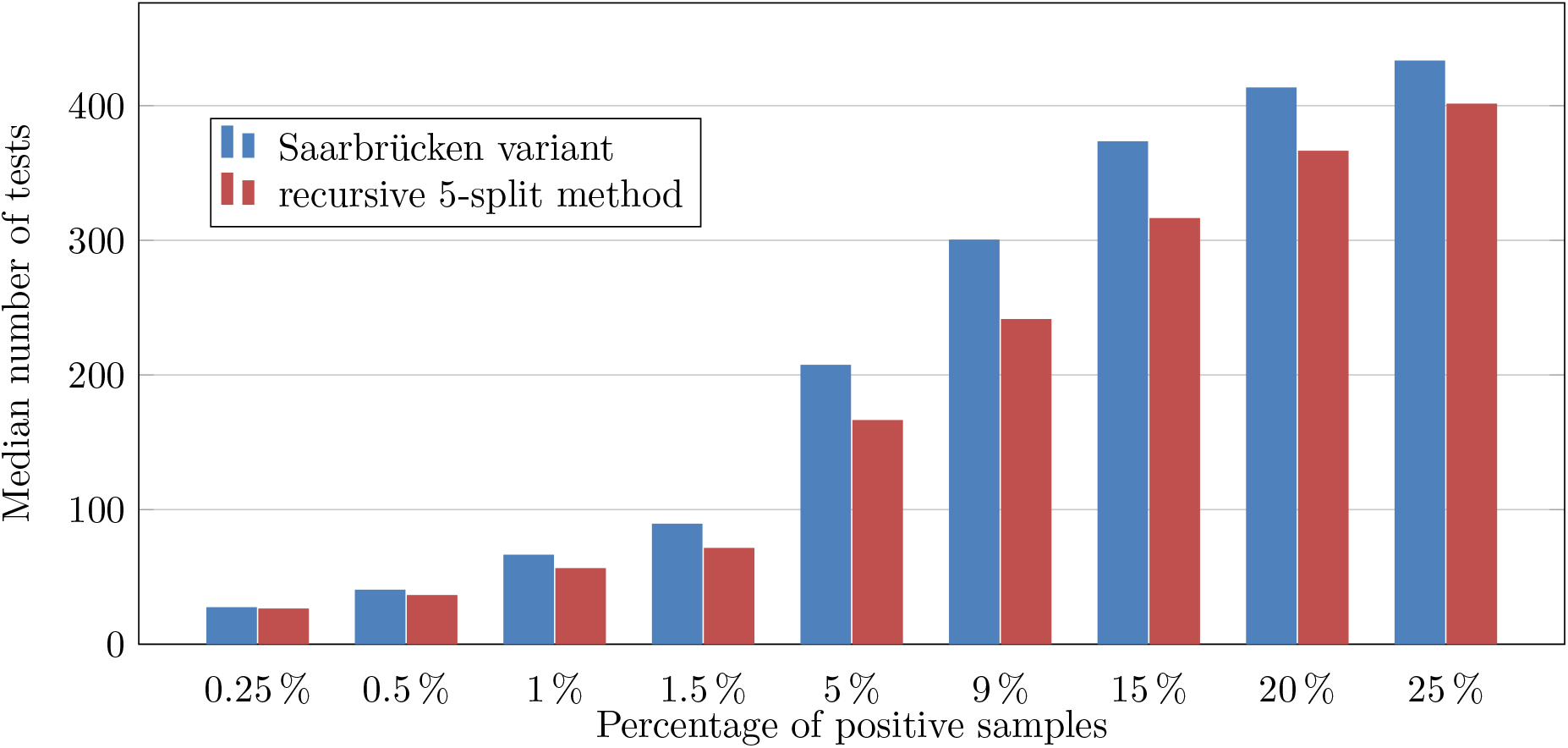
Median number of tests for 400 samples with infection rates from 0.25% to 25%

Compared to current individual testing methods without pools, our experiments show that with a positive sample rate of 1.5 % the number of tests could be reduced by 83.5 %.

**Table 1:**
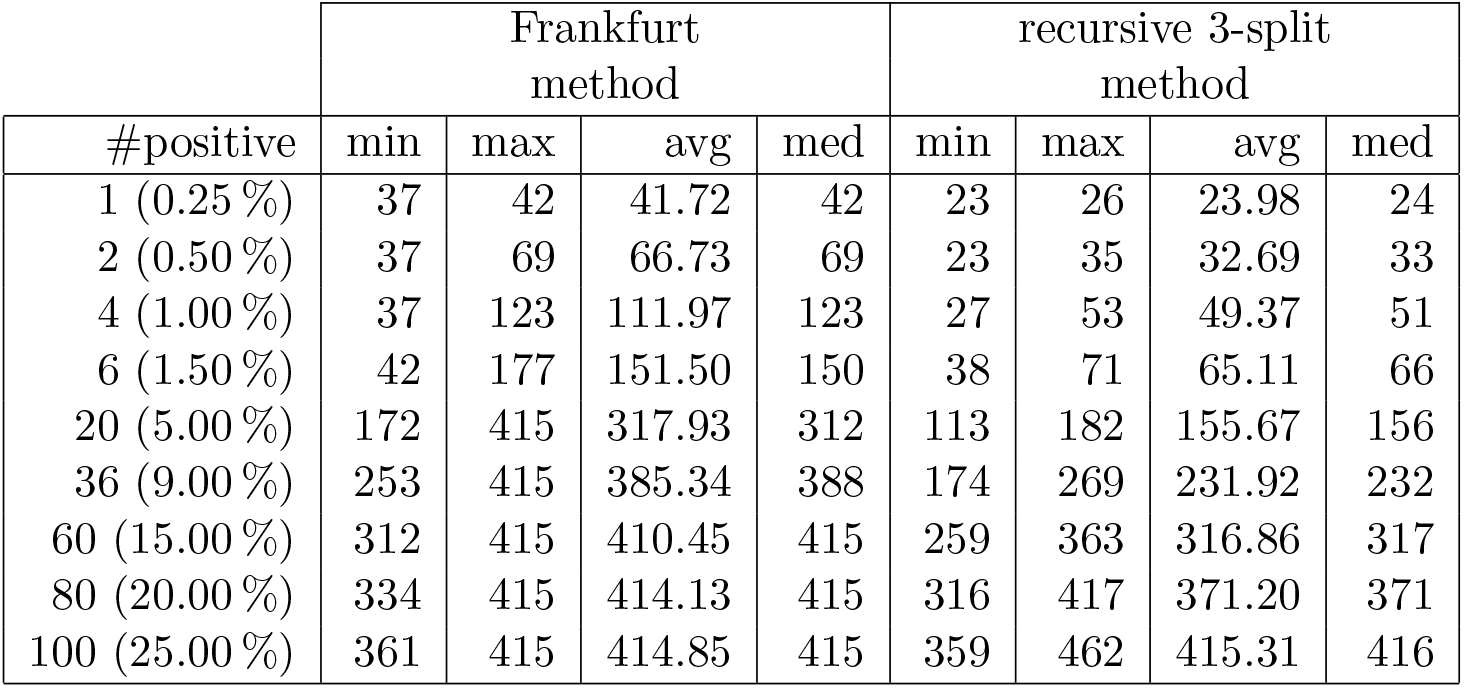
400 samples, pool size 27, statistics over 1,000,000 samples

| #positive | Frankfurt method |  |  |  | recursive 3-split method |  |  |  |
| --- | --- | --- | --- | --- | --- | --- | --- | --- |
|  | min | max | avg | med | min | max | avg | med |
| 1 (0.25 %) | 37 | 42 | 41.72 | 42 | 23 | 26 | 23.98 | 24 |
| 2 (0.50 %) | 37 | 69 | 66.73 | 69 | 23 | 35 | 32.69 | 33 |
| 4 (1.00 %) | 37 | 123 | 111.97 | 123 | 27 | 53 | 49.37 | 51 |
| 6 (1.50 %) | 42 | 177 | 151.50 | 150 | 38 | 71 | 65.11 | 66 |
| 20 (5.00 %) | 172 | 415 | 317.93 | 312 | 113 | 182 | 155.67 | 156 |
| 36 (9.00 %) | 253 | 415 | 385.34 | 388 | 174 | 269 | 231.92 | 232 |
| 60 (15.00 %) | 312 | 415 | 410.45 | 415 | 259 | 363 | 316.86 | 317 |
| 80 (20.00 %) | 334 | 415 | 414.13 | 415 | 316 | 417 | 371.20 | 371 |
| 100 (25.00 %) | 361 | 415 | 414.85 | 415 | 359 | 462 | 415.31 | 416 |

## 3 Limitations of pool testing

The Frankfurt pool testing method increases the test capacity significantly and our simulations indicate that recursive pool testing has the potential to increase the test capacity even further by a factor of about 2 in realistic scenarios. However, pool testing in general and the recursive method in particular have drawbacks that limit their applicability. We recall that we refer to the pool size as *p* and to the required number of aliquots as *a*.

### 3.1 Desirable pool sizes

As Lohse et al. [4] state, borderline samples might escape detection in pools of size 30. However, they expect those samples might stem from almost recovered persons.

With increased *p*, sensitivity, i.e., the chance to detect a positive sample in a pool of negative samples, diminishes. In PCR technology, ideal amplification follows a 2*c* pattern where c is the cycle number. Assuming a 40-cycle PCR, one can estimate the amount of amplified DNA by *amplified_DNA* = *initial_DNA* × *240*. Typically, a realtime PCR reliably detects positive samples as long as the target reaches the threshold value by cycle 37 or 38. With each 1:2 dilution (50% virus concentration) the ct-value is increased by 1. So, for a 1:*p* dilution, ct values increase by log_2_ *p*. With a 128-size pool, one can assume an increase in the ct value from a single sample to a pool of log_2_ 128 = 7. This means that samples with single-sample ct values 31 or above might escape detection. With this in mind, the pool size can be determined by counterbalancing cost and sensitivity.

As a working hypothesis for the following, we assume that pools should not be larger than 30, on the other hand, it is desirable to stay close to 30.

### 3.2 Number of required aliquots

Individual testing requires no sample splitting into aliquots, in our notation *a* = 1, the Frankfurt method requires up to *a* = 2, the Saarbrücken variant up to *a* = 3, and our recursive method with 3-splits up to *a* = 1 + log_3_ *p* aliquots of each sample, at pool size *p* = 9 the latter amounts to *a* = 3 aliquots like the Saarbruücken variant, at pool size *p* = 27 the latter amounts to *a* = 4 aliquots. It is unclear, see also below, if *a* = 4 is tolerable in laboratory practice. We have pointed out the superiority of the 3-split recursive method, but we have not yet taken a limitation of the number of required aliquots into account. If *a* ≤ 3 is required, the 3-split method would have to set the pool size to only 9 which is undesirable. Likewise, the 4-split recursive method (*B* = 4) with “natural” pool size *p* = 4^2^ = 16 is undesirable due to small pool size. On the other hand, 6-split with “natural” pool size *p* = 6^2^ = 36 is undesirable, because the pools get too big. An interesting alternative is to consider 5-splits (i.e., take *B* = 5) and set the pool size to *p* = 5^2^ = 25.

We have repeated the experiment reported in Table 1 and Figure 4 with this alternative in order to see if the number of tests deteriorates in comparison to the previous table for which up to 4 aliquots were needed. We also ran the Saarbruücken variant on the same instances in order to see how well it performs in comparison. The results are given in Table 2 and Figure 5. We observe that recursive 5-split consistently outperforms the Saarbrücken variant and its performance only moderately deteriorates in comparison to the 3-split recursive method.

We also compared the same two methods as well as the 3-split recursive method on 1,000,000 random instances of the original Saarbrücken experiment, i.e., *n* = 1,191 samples, 23 of which positive, randomly distributed, 1,000,000 instances. The pool sizes are 30 for the Saarbrücken variant, 25 for the 5-split, and 27 for the 3-split recursive method, see Table 3. The 5-split recursive method significantly outperforms the Saarbrücken method (both require up to 3 aliquots) even though it uses pools of size 25 rather than 30, and its performance is is pretty close to the one of the recursive 3-split method that requires up to 4 aliquots.

**Table 2:** 400 samples, pool sizes 30 for Saarbrücken variant and 25 for recursive 5-split, statistics over 1,000,000 samples

|  | Saarbrücken<br>variant |  |  |  | recursive 5-split<br>method |  |  |  |
| --- | --- | --- | --- | --- | --- | --- | --- | --- |
| #positive | min | max | avg | med | min | max | avg | med |
| 1 (0.25 %) | 24 | 27 | 26.92 | 27 | 26 | 26 | 26.00 | 26 |
| 2 (0.50 %) | 24 | 40 | 39.42 | 40 | 26 | 36 | 35.65 | 36 |
| 4 (1.00 %) | 27 | 66 | 63.15 | 66 | 31 | 56 | 53.97 | 56 |
| 6 (1.50 %) | 37 | 92 | 85.35 | 89 | 41 | 76 | 71.08 | 71 |
| 20 (5.00 %) | 125 | 253 | 207.06 | 207 | 106 | 196 | 165.61 | 166 |
| 36 (9.00 %) | 201 | 383 | 296.93 | 300 | 176 | 276 | 240.00 | 241 |
| 60 (15.00 %) | 284 | 453 | 375.56 | 373 | 256 | 376 | 318.09 | 316 |
| 80 (20.00 %) | 323 | 453 | 411.21 | 413 | 301 | 436 | 365.48 | 366 |
| 100 (25.00 %) | 353 | 453 | 431.30 | 433 | 336 | 461 | 401.80 | 401 |

**Table 3:**
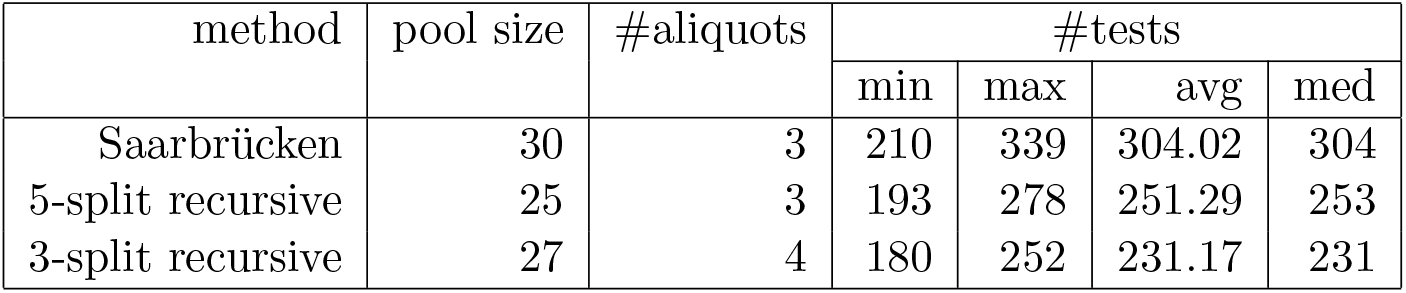
1,191 samples, statistics over 1,000,000 random instances

However, with the 5/25 pooling, a lab can use the same pooling device with identical settings to produce 25-size pools by just using the previously generated size-5-pools as input and still keep the original (aliquot 1) as well as the size-5-pools (aliquot 2) and obtain the 3rd aliquot by making up the 25-size-pool. No re-programming is needed, and lab personnel does not need to keep in mind whether to pool 3, 9 or 27 samples as it would always boil down to 1 in 5.

### 3.3 Required time for all results

With current technology, a batch with up to 94 samples takes a little over 3 hours to be analysed. Any time to pool samples would have to be performed beforehand and laboratory experience indicates that this takes about another roughly 2 hours for 380 samples (pooled into pools of size 5, thus 76 pools). Assuming no invalid results and only valid batches, as well as neglecting setup-times and time taken to produce the pools, individual results are available after 3*a* hours, i.e., 3 hours for individual testing (up to 94 samples), 6 hours with the Frankfurt method, 9 hours with the Saarbrücken variant as well as the 5-split recursive method at pool size 25, and 12 hours with the recursive 3-split method at pool size 27.

While it takes 8 hours to test 1,000 samples using a roche cobas 8800 analyzer, the authors have calculated that with pool size 5 it takes a little more than 14 hours for the same samples (1% positives), and, with recursive pool testing at size 25 and 5, the turnaround-time would go up to 17 hours. While the extended waiting times are intolerable in many scenarios like testing due to individual doctoral prescription or testing in known hot spots, they might well be tolerable in mass testing without previous indications, e.g., when opening a school with 1,000 students and teachers and testing at fortnightly intervals for possible early detection of infections, mainly in settings with very few or no positive samples to be expected.

### 3.4 Laboratory practice

Of course, in order to convince laboratories to undertake changes in their workflow that are prone to prolonged turnaround times, the course of action must be provided in an easily digestible form. The question is: How could something like this be realized in practice?

First and foremost, the LIS (lab information system) needs to be able to distinguish between samples which can be pooled (let’s call them “screening samples”) and samples which must not be pooled (“diagnostic samples”). Those screening samples need to be entered into the system with a specific request, different from the diagnostic samples’ entries. Also, the user (not necessarily the samples’ originators) need to be able to see, at any time, at which point in the prospective pooling step a sample might be just now. Therefore, it might be prudent to allow for 3 different analyses if working with 2 different pools: *“number_analyses* = *number_aliquots”*. We start with aliquots *s_i_*_,1_ of the *n* samples (*s*_1_, *s*_2_, …, *s_n_*} and add *p* samples into one pool which is then analysed using analysis *corona_pool_1*. If *corona_pool_1* turns out negative, the lab order for those patients is fulfilled and the negative result is reported. All those with either a positive or invalid result appear with “not negative” (and hidden to the sender) on the lab report and make the next step (request) appear (open) on the lab report *corona_pool_2*. Same here, any negative pools end up with the negative result behind this analysis while any positive or invalid results appear as “not negative” internally within the lab and make the third analysis appear, visibly, on the report *corona_screen*. All 3 analyses might have the same friendly name appear on the lab report, visible to the samples’ sender.

The strict consecutive pattern of dividing positive pools into subpools described in Section 2.2 has been useful for a concise description of the algorithm, but it is not necessary for the correctness of the recursive method. Rather, different aliquots of the samples in a positive pool can be arbitrarily assigned to the subpools.

Within the lab, samples will be archived and can be retrieved if any pool is positive. In order for the lab personnel to know whether to pool the samples with 26 or 8 or 2 or whatever number of other samples, samples should be stored in different archives and marked with different colours, depending on their status within the workflow chain.

An easier setup would be a pooling device that inherently can pool 5 samples and use this to undergo a 2-step pooling. Step 1: Make pools of 5 of all screening samples. Step 2: Make pools of 5 of all pools from step 1. This way, using the same pipetting robot and the same programming, the complete chain of barcode numbers can be passed on to the LIS after step 2 is complete. This way, also, all containing pools of 5 which might be needed if the 25-size-pool comes back positive, are already available and ready for second stage testing. The downside is: With low prevalence, a significant number of size-5 pools are created unnecessarily, costing time.

However, already having the LIS know the next smaller pools barcode makes it easier to work with residual lists, i.e., which samples still have no result and where do I find them.

## 4 Discussion

These days we face the situation that some restrictions imposed due to the Covid-19 pandemic are reduced stepwise and another shutdown would be disastrous for the economy. We regularly test the players in the German soccer leagues but have no plan how to detect new emerging hotspots in, say, re-opened kindergartens, schools, and retirement homes as early as possible. It seems that a substantially increased testing capability might help in developing an enhanced strategy in dealing with this situation. This view is shared, e.g., by the German Federal Minister of Health Jens Spahn [3]: “Es ist viel teurer, zu wenig zu testen, als zu viel zu testen.”^2^

Currently (end of June 2020), the German Federal State of Bavaria offers preventive SARS-CoV-2 mass testing to all (roughly 13,000,000) state inhabitants. We believe that pool testing in general and the recursive method in particular may be the method of choice.

However, one must not assume that sample pooling reduces the cost of testing by a factor of the pool size. It is clear that laboratories save on reagents with the reduced number of tests. With recursive pooling, we have shown that reagents use can be massively reduced. For positive sample rates of 1.5%, the reduction is about 83% with respect to current individual testing and 56% with respect to the Frankfurt method, respectively. However, one must consider the downsides, namely the increased personnel and other consumable costs (like secondary tubes and pipette tips) due to longer hands-on times and more procedural steps. A major problem could also be the significantly prolonged turnaround times. However, this setup is certainly feasible for laboratories with limited PCR-capacity or countries with limited reagent supplies as well as those having to deal with sample numbers exceeding the laboratory capacities by a large factor over a prolonged period of time. We definitely think that recursive pool testing may be useful in mass testing when only few infections are to be expected.

We can suggest to labs involved in SARS-CoV-2 testing to consider evaluating this setup with their own equipment and calculate the “sweet spot” for their circumstances based on an arbitrary number of samples with varying positivity rates.

While it is also possible to transfer this idea to antibody-testing, one needs to keep in mind that antibody tests mostly follow a linear dilution formula while the exponential increase in DNA-fragments in PCR technology allows for greater dilution factors with relatively smaller impact on possible false-negatives.

## Data Availability

We only use randomly generated data for our experiments. The generation of this data as well as the implementation of our code is explained in the paper, therefore reproducibility is guaranteed.

## 5 Acknowledgments

We gratefully acknowledge the advice of Ulrich Jürgens, Andrea Krieger, Dirk Schmidt, and Norbert Schongen.

## A Appendix: Optimum Split Numbers

We derive the (expected) optimum split numbers *B* for the cases that in a pool of size p there is exactly one (see A.1) or there are exactly two (see A.2) positive samples, respectively, as claimed in Section 2.4. We have *B* ≥ 2 and *B* ≤ *p*. In order to simplify the mathematics we assume *p* to be a power of *B*, i.e., *p* = *B^k^* for some *k* ∊ ℕ.

The set of all possible tests is illustrated by the split tree. Figure 6 displays the split tree for *B* = 3 and pool size *p* = 27. Each box shows a pool with its size (the number of samples in this pool) provided in the box. The split tree has exactly log*_B_ p* + 1 levels, we will denote them by level 0 (the root level), followed by the levels 1,2,…, log*_B_ p*. On the first level *j* = 0 of the split tree all samples are part of the pool of size 27. On the second level (*j* = 1), the samples have been separated into *B* = 3 pools of size *p/B* = 9. The last level (*j* = log*_B_ p*) contains pools with singletons only.

**Figure 6:**
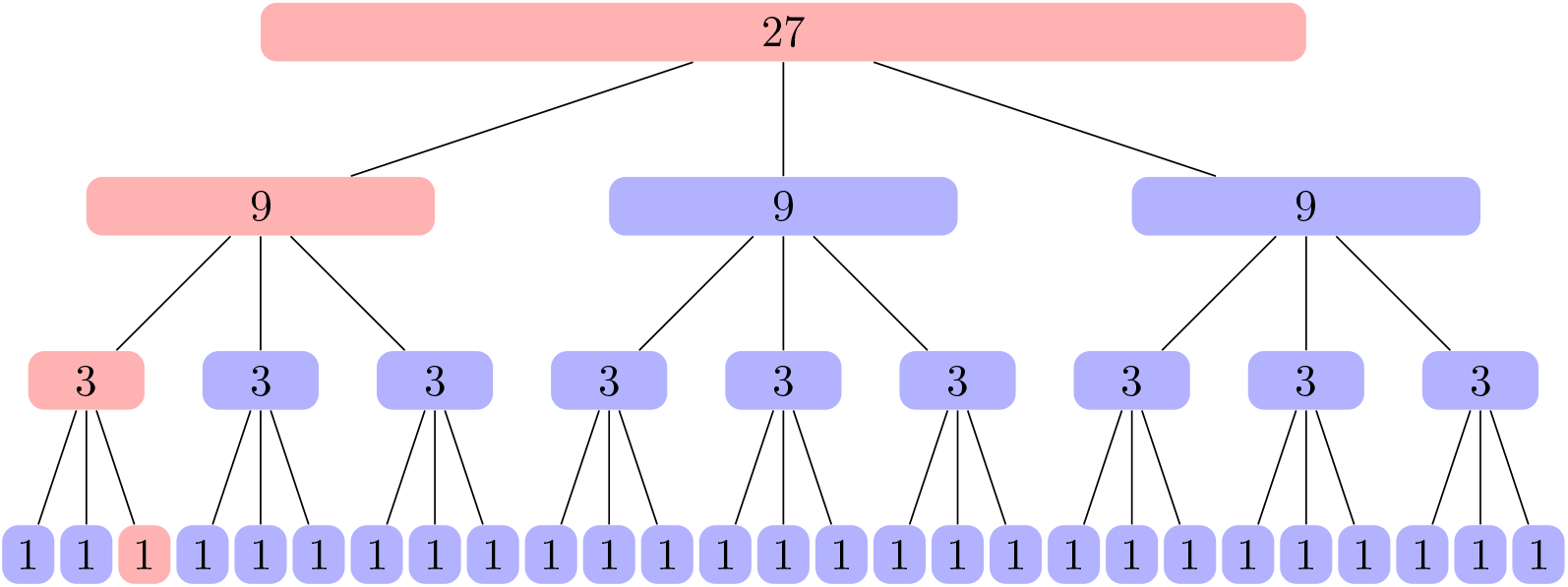
The split tree for *B* = 3 and pool size *p* = 27. Each box corresponds to a pool whose size is given in the box. The red boxes indicate positive pools under the assumption that here is exactly one positive sample. Regardless of the position of the positive sample (here position 3) there is always one positive pool on each level such that a total of 1 + *B* log*_B_ p* = 10 tests are needed.

In general all samples on level 0 are within the same pool of size *p*, on level 1 we have *B* pools of size *p/B* each, on level 2 we have *B*^2^ pools of size *p/B*^2^ each, on level log*_B_ p* − 1 we have *p/B* pools of size *B*, and on the last level we have singletons only.

### A.1 Case 1: One positive sample in the pool

In the case that exactly one sample in the pool is positive, it is obvious that we need one test for the original pool of size *p* (level *j* = 0 of the split tree) and *B* tests on each level of the split tree in order to identify the sample. Hence, for *p* ≥ 2, the number of tests is provided by the function

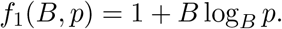

Calculating the first partial derivative with respect to *B*

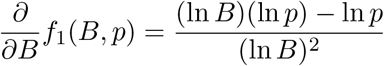

and setting it to 0 gives the minimum *B* = *e* = 2.718…, where e is the Euler number. However, we need to restrict *B* to be integer. We can easily see that *f*_1_(2,*p*) > *f*_1_(3,*p*) for *p* ≥ 3, and *f*_1_ is monotonically increasing with increasing *B* > *e*, therefore we obtain *B* = 3 as the optimum value for *B*. Notice that the function *f*_1_(*B*,*p*) is the same in the best possible case, the worst possible case, and the (expected) average case.

### A.2 Case 2: Two positive samples in the pool

In the case that there are exactly two positive samples in the pool of size *p*, the optimum split number depends on their explicit position. If we are lucky, both samples belong to the same pool all the way down in the split tree (they are separated on the last level only). In this (best) case we need the same number of tests as in the subsection before (see A.1), namely

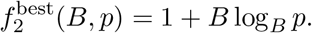

On the other hand, in the worst case the two positive samples are in different pools already on level *j* = 1 of our tree (as shown in Figure 7), in which case we need 2*B* tests for levels *j* = 2 to *j* = log*_B_ p* in addition to the test at level *j* = 0 and *B* tests in level *j* = 1. This sums up to the worst case number of tests

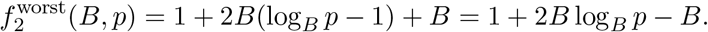

**Figure 7:**
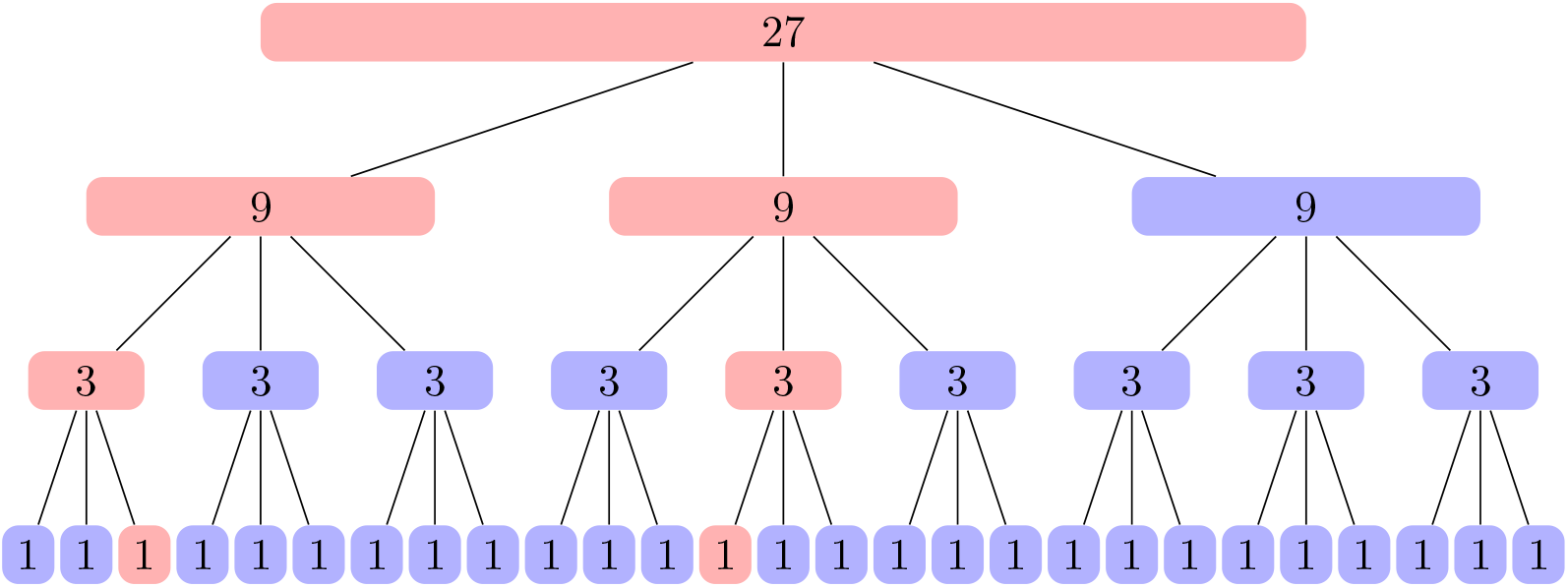
A split tree for *B* = 3 and pool size *p* = 27 with two positive samples. The pools containing positive samples are shown in red. After level *j* = 0 the two samples have been separated into different pools.

In practice, best and worst cases will occur rather rarely. Therefore, in addition to best case and worst case analysis, an *average case analysis* is the appropriate analysis for practical scenarios. In the average case analysis we are interested in the expected number of tests for the following stochastic experiment in which we randomly choose the two positions of the two positive samples in the pool. We introduce the random variable *X* as the number of necessary tests in order to detect the two positive samples correctly. For our analysis we are interested in the expected number of tests **E**[*X*]. We will search for all possible pairs of positive samples once and average over the number of all possible pairs which is 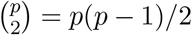.

The number of tests depends on the level of the tree on which the two samples are separated into different pools. Let *j* be defined as the level number for which the two positive samples share the same pool, but are in different pools on level *j* + 1. For the two positive samples (displayed in red) in Figure 7 we can see that *j* = 0. Obviously, all pairs of positive samples satisfying the above condition for the same *j* will need the same number of tests. Let *X_j_* be the random variable of the number of tests needed for those pairs for a specific *j*.

The expected number of tests **E**[*X*] is defined by the sum of all possible outcomes of the random variable *X* weighted by the number of times this outcome is produced divided by the total number of possible pairs. In other words, it is the average number of tests needed to search for all possible pairs:

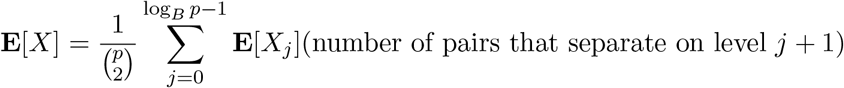

We first consider the number of tests needed when the two positive samples separate directly after level *j*.

- For the latest possible *j* = log*_B_ p* − 1, we have already argued that we get the best case which is 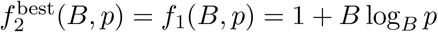.
- For *j* = log*_B_ p* − 2 we need *B* more tests, since the two positive samples are in different pools on level log*_B_ p* − 1.
- For each *j* decreasing by one, we have *B* more tests.
- We know already that for *j* = 0 we get the worst case function 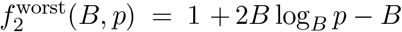.
- Altogether the number of tests in the considered case for *j* is equal to

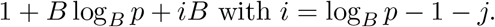 In particular, for *j* = 0 we get 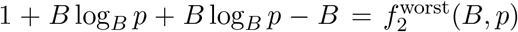 (worst case function) and for *j* = log*_B_ p* − 1 we get 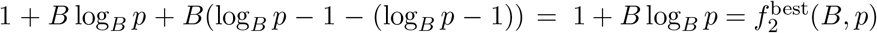 (best case function). Next we calculate the number of pairs that separate directly after level *j*:
- For the latest possible *j* = log*_B_ p* − 1 we have 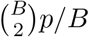 possibilities to choose the two positive samples. In this case both samples are in the same pool until the end. The 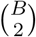 is the number of possibilities for choosing the two samples from the last pool which has size *B*. On the latest possible level we have *p/B* such pools.
- For level *j* = log*_B_ p* − 2 we have 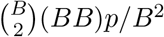 such pairs. Again, we have 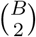 different possibilities to choose the two samples from the last pool which has size *B*. Now for each subtree we need to decide in which of the *B* × *B* positions each of the two samples will sit. Hence, these are (*BB*) possibilities. We have *p/B*^2^ such subtrees rooted at level log*_B_ p* − 2.
- For level *j* = log*_B_ p* − 3 we have 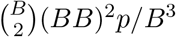 such pairs. Again, we have 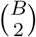 different possibilities to choose the two samples from the last pool which has size *B*. Now for each subtree we need to decide in which of the *B* positions each of the two samples will sit. Hence, these are (*BB*)^2^ possibilities. We have *p/B*^3^ such subtrees rooted at level log*_B_ p* − 3.
- ⋮
- For level *j* = 1 we have 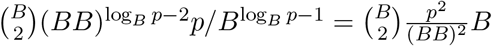 such pairs. Notice that 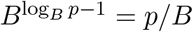 and 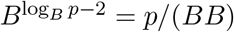.
- For level *j* = 0 we have 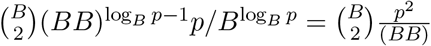 such pairs.
- Altogether for general *j* with *j* = 0,…, log*_B_ p* − 1 the number of pairs separating directly after level *j* is given by

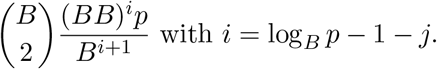 In particular, for *j* = 0 we get 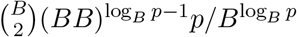 and for *j* = log*_B_ p* − 1 we get 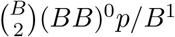.

Putting this together leads to the following function for the expected number of tests depending on *B* and *p* with *B* < *p* and *p* = *B^k^* for some integer *k*:

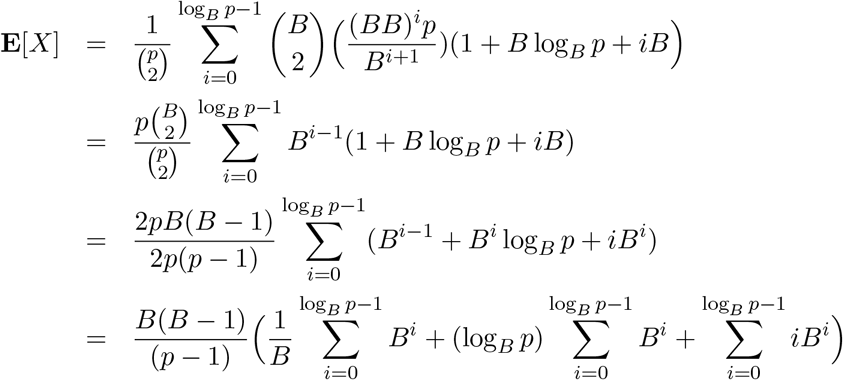

In order to calculate the sums, we use the well known formulae for geometric series

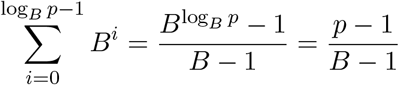

and

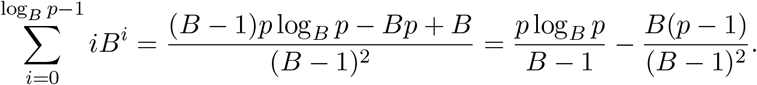

We obtain:

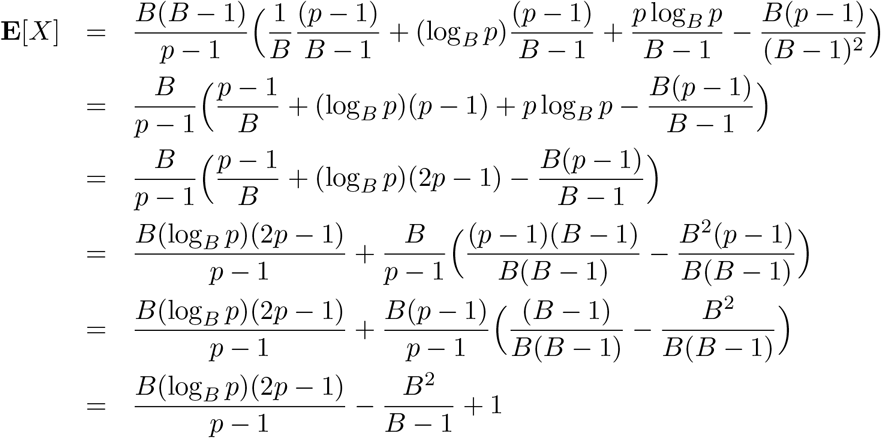

The expected number of tests depends on *B* and *b*, so it can be written as a function *f*_2_(*B*, *b*). In contrast to the case of only one positive sample in the pool, the minimum of the function *f*_2_(*B, b*) depends on the pool size *p*.

For pool size *p* = 6, the best choice is *B* = 5 since the minimum of the function is taken for value 5.284…. For smaller pool sizes *p* ≤ 5, it is preferable to test individually. For pool sizes 7 ≤ *p* ≤ 12, the best choice is *B* = 4 since for these pool sizes the minimum values are within the range [3.507, 4.295], and for pool size *p* ≥ 13, *B* = 3 is the optimum split value: For *p* = 13, the minimum is taken at 3.455… and for larger pool sizes *p* > 13 the minimum again converges to the Euler number *e* = 2.718…. However, this convergence is quite slow. Whereas for *p* = 30, the minimum value of the function is 3.158…, for the (unrealistic) large *p* = 1,000,000, the minimum value is 2.790… .

## Footnotes

1 “Here the swab is first put into an archive tube and subsequently into a pool container. Since this pool method does not increase the volume in the pool container, no dilution and therefore, no loss of sensitivity is observed.”

2 “It is much more expensive to test too little than it is to test too much.”

## Notes

### Competing Interest Statement

The authors have declared no competing interest.

### Funding Statement

No external funding for the work described in the manuscript has been obtained.

### Author Declarations

We do not use any patient data at all.

### Summary of Updates

We have added an Appendix that contains the mathematical analysis.

